# The role of natural language processing in cancer care: a systematic scoping review with narrative synthesis

**DOI:** 10.1101/2024.11.18.24317479

**Authors:** Mengxuan Sun, Ehud Reiter, Lisa Duncan, Rosalind Adam

## Abstract

**Objectives:** To review studies of Natural Language Processing (NLP) systems that assist in cancer care, explore use cases and summarize current research progress.

**Methods:** A systematic scoping review, searching six databases (1) MEDLINE, (2) Embase, (3) IEEE Xplore, (4) ACM Digital Library, (5) Web of Science, and (6) ACL Anthology. Studies were included that reported NLP systems that had been used to improve cancer management by patients or clinicians. Studies were synthesized descriptively and using content analysis.

**Results:** Twenty-nine studies were included. Studies mainly applied NLP in mixed cancer types (n=10, 34.48%) and breast cancer (n=8, 27.59%). NLP was used in four main ways: (1) to support patient education and self-management; (2) to improve efficiency in clinical care by summarizing, extracting, and categorizing data, and supporting record-keeping; (3) to support prevention and early detection of patient problems or cancer recurrence; and (4) to improve cancer treatment by supporting clinicians to make evidence-based treatment decisions. Studies highlighted a wide variety of use cases for NLP technologies in cancer care. However, few technologies had been evaluated within clinical settings, none were evaluated against clinical outcomes, and none had been implemented into clinical care.

**Conclusion:** NLP has the potential to improve cancer care via several mechanisms, including information extraction and classification, which could enable automation and personalization of care processes. Additionally, NLP tools such as chatbots show promise in improving patient communication and support. However, there are deficiencies in the evaluation and clinical integration challenges. Interdisciplinary collaboration between computer scientists and clinicians will be essential if NLP technologies are to fulfil their potential to improve patient experience and outcomes. Registered Protocol: https://doi.org/10.17605/OSF.IO/G9DSR

## 1. Introduction

Cancer is a major public health problem that leads to premature deaths worldwide [1, 2]. Cancer care includes various clinical processes, from screening and early detection, through treatment, survivorship care, and end-of-life care [3].

Improvements in cancer care processes require both technical innovation and medical expertise. Natural Language Processing (NLP) is a branch of Artificial Intelligence (AI) that focuses on the recognition, understanding, and generation of text. It merges linguistics and computer science (e.g. machine learning and deep learning) methodologies.

NLP could be used to enhance the efficiency and quality of cancer care and to improve patient experience within cancer care [4, 5], but the full potential of this emerging technology in oncology is not yet clear [6].

Potential applications include chatbot assistants [7], systems that summarise complex documents [8] or extract meaningful information from a large amount of data [9], and systems that can recommend articles [10] or make treatment suggestions [11]. In the context of cancer care, NLP could play a significant role in analysing unstructured text data, turning multi-source and complex information into an easily understandable format and sharing clinical knowledge.

In cancer care, text data are generated from various sources, including electronic health records (EHRs), diagnostic reports, and patients’ self-reports (for example, via online forums or patient-reported outcome monitoring) [12]. It is possible that NLP could be applied to this data to improve clinical care processes and ultimately, to improve outcomes for patients.

Despite the breakthroughs in technology, a gap exists in the integration of NLP into clinical workflows. Challenges to this integration include data privacy concerns, clinical validation, and the translation of algorithmic outputs into actionable insights [13]. The scope, challenges, and complexities of using NLP potential to improve the efficiency and effectiveness of cancer care have not been formally evaluated.

We conducted a systematic scoping review to investigate the role of Natural Language Processing (NLP) in cancer care. The aim was to explore applications of NLP in cancer care, to summarize the NLP methods used, to analyse the results of evaluations of these technologies, and to identify potential limitations of the systems and their evaluations. This research aims to provide a comprehensive overview of the specific applications of NLP in cancer care, to guide future policy and research.

## 2. Methods

A systematic scoping review was conducted to examine the literature, and “the extent, range, and nature of research activity” of NLP research in cancer care [14]. Methods encompassed descriptive and narrative synthesis to gain a more comprehensive understanding of the research landscape and gaps in this field.

The scoping review approach was chosen because NLP is a potentially broad and complex domain, and the full characterization of the literature involves disparate use cases and technological approaches. Moreover, due to the rapid advancement of NLP technologies and heterogeneity in intended outcomes in the medical domain, it may be challenging to find formal clinical trials for quality assessment. Therefore, a scoping review for this domain is better suited than a traditional systematic review.

We did not interact with the NLP systems directly or test their effectiveness in real-world use cases. Considering the fast-paced development of NLP technology, we aimed to include as many relevant studies as possible and to categorize the way that NLP has been applied in cancer care. These categories will be relevant, even as technical improvements to NLP improve the effectiveness of the technology. A protocol S3 File presented the research design.

### 2.1 Database search strategy

Keywords were selected, truncation symbols and wildcards were combined with Boolean operators to search databases encompassing medical, NLP, engineering, and scientific sources. Databases included six platforms: (1) MEDLINE, (2) Embase, (3) IEEE Xplore, (4) ACM Digital Library, (5) Web of Science, and (6) ACL Anthology. MEDLINE and Embase are widely used for biomedical research. IEEE Xplore and ACM Digital Library are comprehensive full-text repositories of high-quality engineering, computer science, and technology literature.

Relevant Medical Subject Headings (MeSH) were selected, and synonyms were considered to broaden the scope. The database search strategy was formulated in consultation with a senior medical research librarian. The publication time is from 2013 to January 2023. The full database search strategies are presented in S1 File.

### 2.2 Inclusion and exclusion

Inclusion and exclusion criteria are shown in Table 1. Studies were included where NLP had been applied to improve cancer care by patients or clinicians. All cancer types were included, and there were no age restrictions.

**Table 1.** Inclusion and exclusion criteria.

| **Criteria** | **Inclusion** | **Exclusion** |
| --- | --- | --- |
| **Focus of Studies** | Applying NLP to enhance cancer management | Not centred on both cancer management and NLP |
| **Cancer Types and Stages** | Any cancer type and stage after diagnosis | Studies where there is not a focus on confirmed cancer diagnosis.<br>Predictive technologies, cancer risk, prevalence, or assessment for potential symptoms, death-related applications |
| **Participant Inclusion** | All ages - children, adults; NLP for clinical care | Use cancer datasets for NLP research without focusing on improving cancer management by patients, caregivers, or clinicians. |
| **Technological Scope** | NLP being applied to a clinical use case (to assist patients, carers, or professionals with an aspect of cancer management) | NLP for database searches (e.g., PubMed) |
| **Publication Sources** | Research articles in journals or conferences | Studies without accessible full-text publications or detailed conference abstract descriptions |
| **Study Types** | Clinical trials, development papers, feasibility studies | Studies lacking medical or NLP evaluation |
| **Language/Programming Restrictions** | No restrictions | - |

### 2.3 Data screening

Article screening followed the PRISMA flowchart [15] (Fig 1) with four main steps: (1) Titles and abstracts were independently dual screened by four authors (MS screened all titles, RA/ER/LD screened a third of titles each) 2. Discussion about conflicts: the four authors met to discuss any titles/abstracts for which there was disagreement (3) Full-text screening-the four authors independently screened full texts (MS screened all full texts, RA/ER/LD screened one third of full texts each (4) Meeting to discuss any conflicts and to decide on the final papers.

**Fig 1.**
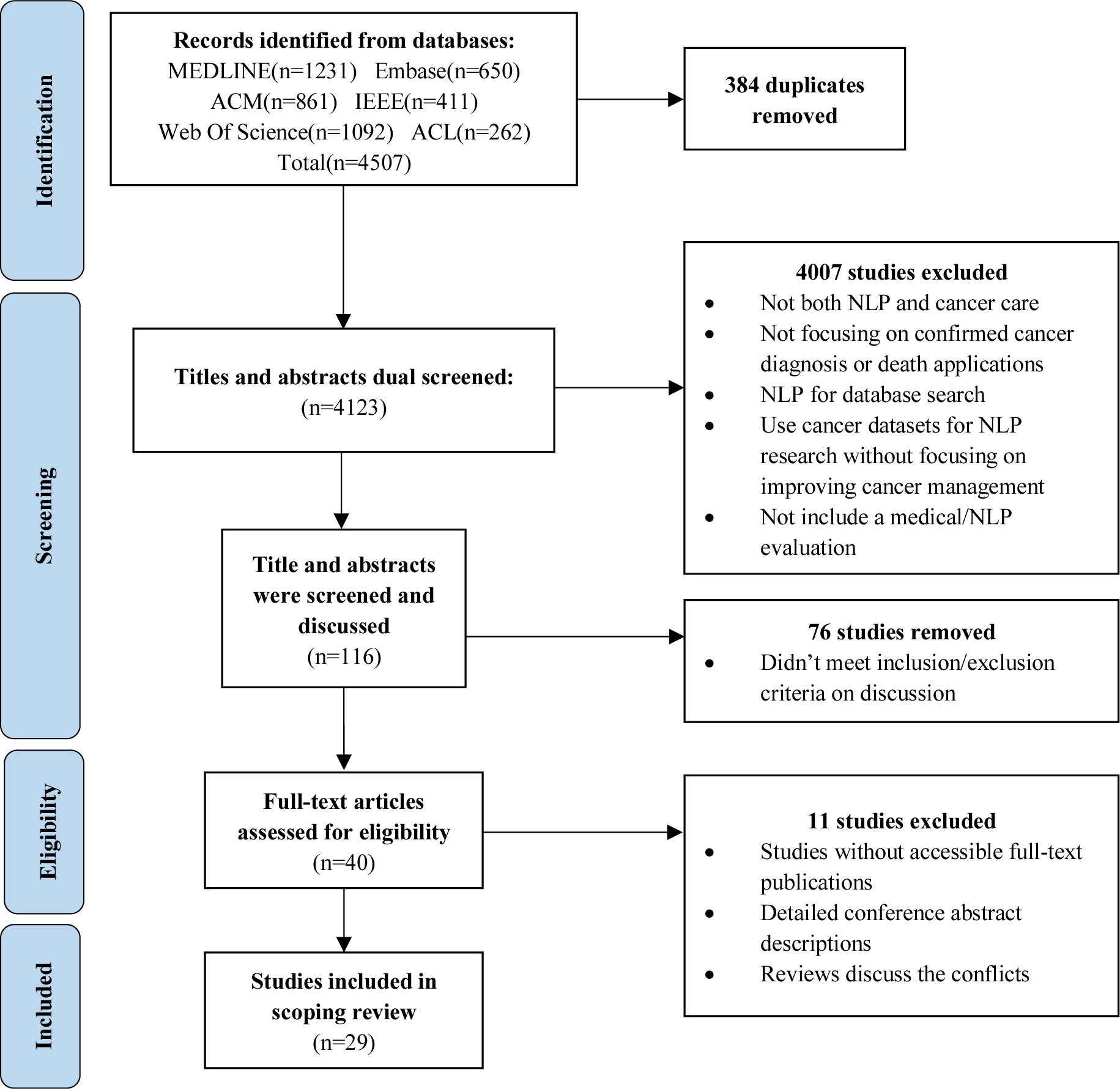
PRISMA Flowchart for study selection.

### 2.4 Synthesis

We synthesized data across the studies using content analysis [16] and descriptive, narrative synthesis. Narrative synthesis and content analysis involved categorizing the content of the studies based on different practical use cases for NLP, considering different experiment data sources, objectives, techniques, clinical issues addressed, results, and evaluations. This approach allowed us to integrate findings from heterogeneous studies leading to a more comprehensive understanding of the scope and context of NLP use in cancer care.

The authors created a bespoke data extraction table in Microsoft Excel. Data were extracted by MS. Extracted data included: Title, Name of the first author, Year, Journal/Conference, Language of applications, Country of origin, General objectives, System Objectives, Cancer type, Distribution of the target audiences, NLP tasks or typical models, Ways NLP were applied in cancer care, Dataset used to train the language model, Evaluation methods, Key findings.

## 3. Results

### 3.1 Description of studies

Table 2 provides an overview of the included studies. Studies were published between 2016 and 2022. The most common country of origin was the United States (n=14, 48.28%) and studies were primarily sourced from medical journals (n=10, 34.48%).

**Table 2.** Included study characteristics.

| **Study characteristic** | **Studies,n,%** |
| --- | --- |
| **Year of publication** |  |
| 2013 – 2016 | 2, (6.9%) |
| 2016 - 2019 | 10, (34.48%) |
| 2020-2023 | 17, (58.62%) |
| **Lead Author's Affiliation** |  |
| United States | 14, (48.28%) |
| Australia | 4, (13.79%) |
| Thailand | 2, (6.90%) |
| Canada | 2, (6.90%) |
| China | 2, (6.90%) |
| Other (Germany,Singapore,Japan,Netherlands,Taiwan) | 5, (17.24%) |
| **Publication type** |  |
| Medical and science journal | 19, (65.51%) |
| NLP conference | 2, (6.90%) |
| ACM | 5, (17.24%) |
| IEEE | 3, (10.34%) |
| **Target Recipients of NLP technology** |  |
| Clinicians | 18, (62.07%) |
| Patients, caregivers, or family members | 8, (27.59%) |
| Online support group therapists | 1, (3.45%) |
| Patients and clinicians | 2, (6.90%) |
| Not mentioned | 1, (3.45%) |
| **Cancer Type** |  |
| Mixed cancer types | 10, (34.48%) |
| Breast Cancer | 8, (27.59%) |
| Lung Cancer | 5, (17.24%) |
| Prostate Cancer | 5, (17.24%) |
| Colorectal Cancer | 4, (13.79%) |
| Other (Primary Cutaneous Melanomas, liver, ovarian, kidney, paediatric oncology, all n=1) | 5, (3.45%) |

The target recipients for the NLP systems were clinicians (n=17, 58.62%). The studies encompassed a wide range of cancer types, with mixed cancer types (n=10, 34.48%) and breast cancer (n=8, 27.59%) being the most frequent focus.

Table 3 illustrates the characteristics from NLP perspective. Electronic medical records (n=11, 37.93%) were the main source of text data, followed by investigation reports (n=5, 17.24%). In terms of NLP tasks, text classification (n=9, 31.03%) and information extraction (n=8, 27.59%) were most common, with NLP principally being used to categorize and extract relevant information from complex medical data. The choice of system architecture varied, with pipeline architecture [17] (n=11, 37.93%) and embedding-based models [18] (n=10, 34.48%) emerging as popular choices for structuring NLP systems.

**Table 3.** Include studies NLP characteristics.

| **Type of data in NLP use case** | **Studies,n,%** |
| --- | --- |
| Electronic medical record | 11, (37.93%) |
| Patient forums/online support groups | 4, (13.79%) |
| Databases of medical and research publications | 3, (10.34%) |
| Diagnostic investigation reports (radiology, pathology) | 5, (17.24%) |
| Multiple sources | 2, (6.90%) |
| Survey/Previous study/Kaggle competition dataset | 4, (13.79%) |
| **NLP tasks** |  |
| Text classification | 9, (31.03%) |
| Information extraction | 8, (27.59%) |
| Chatbot | 3, (10.34%) |
| Decision making system | 3, (10.34%) |
| Emotion analysis and prediction | 2, (6.90%) |
| Report generation | 2, (6.90%) |
| Recommendation system | 2, (6.90%) |
| **System Architecture** |  |
| Rule-based | 5, (17.24%) |
| Pipeline Architecture | 11, (37.93%) |
| Embedding-based Model | 10, (34.48%) |
| Transformer Model | 3, (10.34%) |

### 3.2 Use cases and aims for NLP systems in cancer care

Clinical use cases for NLP in cancer care could be classified into four categories:

- Systems to support patient education and self-management (including emotional support, and self-management of distress) (n=7, 24.1%)
- Systems to improve efficiency in clinical care by summarising data, extracting data, categorising data, and supporting record-keeping (n=12, 41.4%)
- Systems to support prevention and early detection of patient problems or cancer recurrence through risk stratification (n=4, 13.79%)
- Systems to improve cancer treatment by supporting clinicians to make evidence-based treatment decisions (n=6, 20.69%)

Fig 2 summarises the clinical applications of NLP systems in cancer care. Each of the papers and the NLP systems are described in detail in terms of scope, evaluation results, and limitations in S2 Table. Examples are given below.

**Fig 2.**
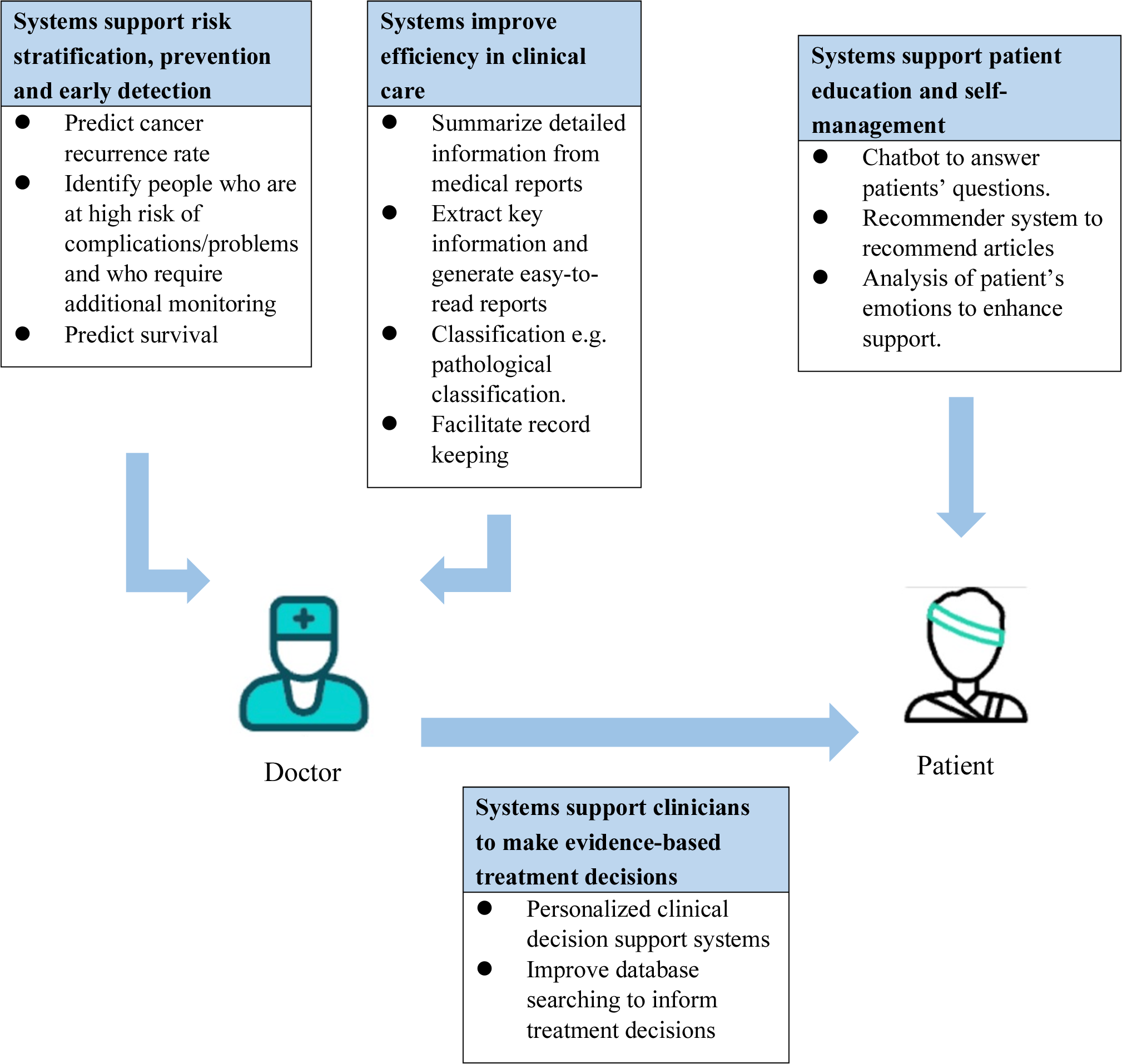
Overview of NLP in cancer care use cases.

#### 3.2.1 Systems to support patient education and self-management (including emotional support, and self-management of distress)

Four studies aimed to use NLP to educate patients [19–22]. A wide variety of NLP technologies were used, including chatbots and systems to recommend relevant articles or information (recommender systems) to patients. Three studies aimed to use NLP for monitoring or analysing patients’ emotions [23–25]. However, most of these studies were exploratory and did not include rigorous evaluation of both NLP model performance and medical user opinions.

### Educating Patients

Kataoka et al [19] developed a web-based chatbot aimed at assisting lung cancer patients and caregivers to manage symptoms. During tests involving medical professionals and patients, the chatbot performed poorly, with 13% of the answers not matching the questions, leading to user dissatisfaction [19].

Hommes et al [20] presented a data-to-text system designed to provide patients with information about quality-of-life implications of cancer treatment to facilitate shared decision-making. Prognosis data was input by the clinician into the prediction model. The system returned messages, such as “X% of patients like you experience difficulties in certain areas, and you may benefit from **” and “50% of patients, like you, face economic issues”. The system was evaluated by two clinicians, who agreed that the messages were useful. However, one physician noted that not all quality-of-life indicators should be conveyed in the same manner. There was no evaluation of system performance.

Chuan et al [21] developed and evaluated a chatbot designed to simplify complex eligibility criteria for cancer clinical trials into easily answerable questions for users. Interacting with the chatbot led participants to become more proactive in asking questions and seeking information.

Rahdari et al [22] built a system to recommend educational information (described as a knowledge-adaptive Interactive Recommender System for Ovarian Cancer Patients and Caregivers, named HELPeR). The system was not evaluated for accuracy or clinical utility.

### Analysing patients’ emotions

Adikari et al [23] developed a chatbot framework to track patients’ emotions in real-time and to provide patient feedback through empathic conversational agents. The model provided text responses and suggested resources and alerts based on patient behavioural metrics. The model achieved a 79% accuracy in correctly predicting positive or negative emotions in the next interaction and 63% in predicting specific emotions among all emotions.

Leung et al [24] developed and evaluated an artificial intelligence–based co-facilitator system(AICF) to help therapists identify participants’ distress through real-time analysis of text-based messages posted during synchronous clinical sessions. AICF successfully extracted emotions and achieved accuracy rates of moderate distress (63%), high distress (85%), and low distress (78%) [24]. The recommendation system equipped in AICF was evaluated by Leung et al [25] based on the ability to appropriately identify keywords indicating psychosocial concerns and make recommendations, aimed to recommend self-management resources to patients. The F1 score (the harmonic mean of precision and recall, a measure of a model’s accuracy) for patient recommendations was 0.880, and 19 of those patients (75%) found the resources useful.

#### 3.2.2 Systems to improve efficiency in clinical care by summarising data, extracting data, categorising data, and supporting record-keeping

Four papers [8, 26–28] summarized detailed information from medical reports for clinicians. Three papers [29–31] used extracted information to generate easy-to-read reports. Four papers [32–35] were designed to classify cancer information. One [36] designed NLP systems to facilitate record keeping (see S2 Table).

### Extracting information from medical reports/records

Mitchell et al [26] fine-tuned a Bert model on pathology reports from a cancer centre (Moffit Cancer Centre, USA) and focused on answering questions from oncology pathology reports. The system was also used to predict cancer site and histology codes according to the International Classification of Oncological Diseases Version 3.2 (ICD-O-3.2) [37]. The model accuracies of questions and answers, site and histology were 79.99%, 73.84%, and 85.29% respectively. Compared to previous research, this study included more cancer categories and obtained higher accuracy scores [37].

Hernandez-Boussard et al [27] focused on developing and validating an NLP pipeline system to assist clinicians in extracting patient-centred outcomes (PCOs) and making classifications to make informed treatment decisions for localized prostate cancer detection. The study was conducted to identify and extract cases and controls of urinary incontinence and erectile dysfunction from electronic health records. Classification results achieved F1 scores of 87% affirmed, 96% negated, and 91% discuss risk(need to discuss the potential risk) for urinary incontinence, 85% affirmed, 92%, negated and 90% discuss risk for erectile dysfunction.

Gupta et al [28] developed a model to generate medical report annotations in clinical practice systems automatically, using NLP to read radiological reports for oncologists and mark the staging of lung cancer. Rule-based system accuracy achieved up to 59% for Tumour (T)-staging, 36% for Node (N)-staging and 41% for Metastasis (M)-staging but their Long Short-Term Memory (LSTM)-based system reached 85% accuracy for the collective TNM staging. Importantly, only 23 annotated reports were used in the evaluation, and the results may not be reliable.

Kim et al [8] applied an NLP program, the Kaiser Permanente Southern California (KPSC) Clinical Information Extraction System, to automatically extract cancer pathology details such as TNM stage, Gleason grade, and tertiary Gleason pattern presence from prostatectomy reports within an electronic medical record system. The system achieved an overall accuracy of 98.7%.

### Restructuring medical reports

Ou et al [29] developed an automated system that extracted summary information from free-text pathology reports to automatically populate structured reports. The system extracted data from 28 categories such as cell growth and microsatellites, and the NLP system combined supervised machine learning CRF with rule-based methods. The overall F1 score was 84.79%. The study wasn’t successful in recruiting pathologists to evaluate the accuracy of the structured outputs.

Savova et al [30] constructed a system named DeepPhe to extract mentions from patients’ radiology and surgical pathology reports, generating summaries of tumour episodes and phenotypes. The system’s performance was compared to human experts’ abstracted information. The agreement between the two human experts ranged from 0.46 to 1.00, and system agreement with humans ranged from 0.20 to 0.96. Thus, the authors asserted that DeepPhe performed similarly to human experts.

In another study, Chuang et al [31] implemented a system that used keyword extraction and machine learning classifiers to analyse unstructured CT reports of liver tumours. The experiments showed that with accurate keywords, the proposed model achieved an AUC (Area Under the Curve, a performance metric commonly used to evaluate the quality of a classifier in binary classification tasks) of 0.95, outperforming other Bert-based pre-trained models [31].

### Classifying data from diagnostic and clinical reports

Four deep-learning-based classifiers [32–35] were designed to perform classification tasks based on medical reports and notes.

Park et al [32] conducted hierarchical cancer-to-cancer transfer and zero-shot string similarity methods, aimed at sharing information to models between different cancers to allow the ability to classify accurately with the minimum labelled data and auxiliary class features in pathology reports. These methods improved the classification performance even with minimal labelled data, achieving a macro F1 score (average performance of each class) of 0.5 and a micro F1 score (classification performance of each sample) of 0.86 (which might be because the data were imbalanced). The authors suggested that pre-training the model on a larger volume of text would improve the results.

Chen et al [33] developed a knowledge-powered deep breast tumour classifier (KDBTC) to simultaneously process multiple medical reports, including Breast-ultrasound, Mammography (X-ray), and Nuclear Magnetic Resonance Imaging (MRI). The goal was to improve the accuracy and coverage of benign versus malignant breast image classification. By combining both the syntax and semantic representations of medical reports, the KDBTC model performed the best with 0.8854 accuracy and 0.9070 F1 score.

Kehl et al [34] developed a method for extracting structured clinical outcomes from unstructured medical oncologist notes. A rule-based classifier was designed to recognize the assessment/plan for a given note, and a Recurrent neural network model (RNN) was trained to predict whether each remaining word in each note was part of the plan. A Convolutional neural network (CNN) model was trained to predict the labels applied by a human curator to assess text for the following three binary outcomes: whether the cancer was present; whether the cancer was progressing or worsening; and whether the cancer was responding to treatment/improving. The results of the method reach AUROCs (Area Under the Receiver Operating Characteristic curve) of 0.94 for the presence of cancer outcome, 0.86 for the cancer progression outcome, and 0.90 for the response to treatment outcome.

Bozkurt et al [35] aimed to classify the severity of urinary incontinence experienced by patients by using a using hybrid NLP pipeline that combined rule-based and deep-learning CNN models, to classify clinical records. The system classified positive urinary incontinence cases as mild, moderate, and severe. The evaluation applied manually annotated data, with F1 scores of 0.76, 0.81, and 0.71 respectively.

### Supporting record keeping

Banerjee et al [36] proposed a weakly supervised NLP method for annotating electronic clinical notes automatically and providing Patient Centred Outcomes for clinicians with insufficient data. The model achieved an average F1 score of 0.86 on the sentence-level three-tier classification task (Presence/Absence/Risk) in urinary incontinence and bowel dysfunction. The authors noted that their system outperformed existing rule-based urinary incontinence annotation models [36].

#### 3.2.3 Systems to support prevention and early detection of patient problems or cancer recurrence through risk stratification

Four articles concerned risk stratification in people with cancer: two [38, 39] predicted the probability of cancer recurrence, one [40] monitored the daily activities of patients and gave an early warning of the need for more frequent patient monitoring, and one [41] predicted survival time.

### Risk assessment and follow-up care monitoring

He et al [40] introduced a mHealth framework that extracts information from medical records and inputs it into a risk assessment calculator to determine whether people who were treated surgically for cancer were at high risk of surgical complications (including hospital readmission, infection, and other risks associated with nutrition and body weight). Those at high risk were monitored more closely using a smartphone app. The app shows a visual display of physical measurement results. The Bayesian information retrieval risk prediction model achieves an accuracy of 97% and a sensitivity close to 100%, but the specificity is only 70%, meaning that a small number of low-risk patients might be misclassified as high-risk, and they could be subjected to unnecessary monitoring.

### Cancer recurrence prediction

Ho et al [38] designed a deep learning architecture to exploit multi-modal healthcare data to predict colorectal cancer recurrence. The system involved a modified Transformer model to extract high-quality features and a multilayer perceptron for learning tabular data features. The model achieved an AUROC score of 0.95, with precision, sensitivity, and specificity scores of 0.83, 0.80, and 0.96, respectively.

Sanyal et al [39] developed a weakly supervised breast cancer recurrence prediction framework. This allows a deep learning model to learn from larger datasets using manually annotated data and system-generated imperfect data to reach better performance. The model achieved an AUROC of 0.94 on validation data from 224 recurrent patients.

### Short-term life expectancy prediction

Banerjee et al [41] proposed the Probabilistic Prognostic Estimate of Survival in Metastatic Cancer Patients (PPES-Met) for estimating short-term life expectancy (>3 months) by analysing free-text clinical notes in the electronic medical record while maintaining the temporal visit sequence, achieving an AUC of 0.89. However, the model lacked transparency and the authors were not able to explain why or how the computer model predicted specific outcomes at particular time points.

#### 3.2.4 Systems to improve cancer treatment by supporting clinicians to make evidence-based treatment decisions

Four articles [42–45] developed personalized clinical decision support systems and compared them to actual treatment decisions. Another two studies [46, 47] aimed to improve clinical decision support by searching in real-world case databases (46), or in PubMed and clinical trials databases (47).

Suwanvecho et al [42] obtained evidence-informed therapeutic options from IBM Watson for Oncology (WfO) and compared them with the actual treatment decisions. The statistical analysis revealed that 60% of the therapeutic options were in line with the actual treatment decisions, 70% involved similar or acceptable alternative treatment options, and 30% were considered unacceptable.

Mokashi et al [43] developed a platform to extract features from online forums to analyse the social needs of breast cancer patients, built connections between patients and topics, to help clinicians estimate non-medical issues for making a personalised treatment. The results showed that the Patient-Topic Network coherence, measured by Normalized Pointwise Mutual Information (NPMI), was 0.481, and the Precision and Recall were 0.643 and 0.588, respectively, surpassing the performance of the word2vec model.

Silva et al [44] developed a framework to aggregate patient information and interaction trajectories from a prostate cancer Online support group (OSG) and studied the influences of online social interactions on treatment type selection, post-treatment recovery, side effects, and emotions expressed over time for different patients.

Becker et al [45] developed an NLP pipeline system to extract unstructured information in colorectal cancer patients’ EHR records and map Unified Medical Language System (UMLS) concepts to identify 11 clinical guidelines indicators such as tumour stage to assist in decision-making. The system was evaluated using data obtained from German EHR clinical notes annotated by a physician. The average precision value across all treatment decision indicators for patients with known cancer diagnoses achieved an F1 score of 81.81%.

Shen et al. [46] utilized triplet information (method, cure, cancer) in the form of ontologies to enhance the effectiveness of Decision Support Systems (DSS). This approach improved knowledge integration and interconnectivity, leading to increased accuracy and reasoning ability when searching for cancer treatment decisions in Case-Based Reasoning (CBR) databases. The evaluation of the ontologies yielded an AUC of 0.846, which surpassed the performance of the systems without ontologies, which was only 0.73.

Koopman et al [47] developed a search engine to assist clinicians in finding targeted treatments for children with cancer by searching PubMed articles and clinical trials. The system map free text to biomedical entities, with a focus on (gene, drug, cancer) relevant to clinicians. The output entities outperformed the information in a knowledge graph and addressed the needs of clinicians. However, the authors did not evaluate the accuracy and usability of the system.

## 4. Discussion

Natural Language Processing has been used in a wide range of settings and clinical use cases to support cancer care. These include supporting patient education and self-management; extracting, summarising and classifying data from clinical reports such as diagnostic reports, and electronic health records; supporting or predicting early detection of patient problems or cancer recurrence; and supporting clinicians to make evidence-based treatment decisions.

### 4.1 Performance and Limitations of NLP technologies in Cancer Care

The NLP systems identified and their evaluations are heterogeneous. We have identified several important considerations relating to NLP performance and potential limitations of the technologies/their evaluations:

#### NLP technology

Most of the technologies in these published reports are several years old. More effective models like BERT or Transformer which were published in 2018 have not been widely used.

There was not always transparency about how models produced their results[41].

#### Data

There was limited availability of private data. Challenges in data annotation can make it difficult to obtain large datasets for training neural network models. [28].

Additionally, incomplete medical cases led to suboptimal accuracy in some studies.

#### Performance

Studies measure the system performance with different indicators, such as accuracy, F1 score, AUC, and AUROC. Many studies that reported accuracy achieve a theoretical score above 80%[8, 27, 29, 31]. However, in medicine, small errors in the accuracy of outputs and more subtle issues with the nature of the language used (e.g. emotionally insensitive responses) could cause significant problems. Furthermore, a few NLP systems [19, 43] only exhibited accuracy as low as 60%, making them insufficient to provide effective assistance to healthcare professionals. NLP systems and tasks that were focused on patients’ lives and feelings were often not evaluated quantitatively or assessed for accuracy at all.[20, 40].

#### Evaluation

Very few NLP studies were evaluated in clinical settings. Testing model performance statistically and in terms of real-world clinical performance are both necessary. In some of the studies in this review, NLP researchers had difficulties recruiting clinicians to perform human evaluation of the system outputs.[28, 40].

#### Robustness (generalizability and coverage)

Due to the wide use of private data and rule-based systems, the proposed methods may not suit the same use case in different data and environments. NLP outputs may lack local context or miss the nuance that is required for clinical decision-making.

#### Implementation

In our review, the technologies evaluated were some way from being implemented directly into clinical care. Whether NLP results can meet medical regulatory standards and how NLP can assist in diagnosis are evolving areas of interest.

In addition, the safety of using AI and whether there is bias [48–50] are also important factors to consider.

### 4.2 Methodological considerations

The scoping review systematically identified an extensive range of studies from both computing science and medical databases. A wide range of use cases for NLP were summarised. Limitations of scoping reviews include that quality assessments of the included articles are not conducted, and there is a certain degree of subjectivity in the selection of literature.

The latest question-and-answer generative large language model ChatGPT [51] is widely used by the public and has implications for many industries, including medicine. Our search was carried out in January 2023, and the model had just been released. The release of ChatGPT will likely result in a proliferation of reports and use cases relating to cancer. Large language models such as ChatGPT will likely be used by both clinicians and patients for the tasks highlighted in this review (see Figure 2), such as summarising complex information and concepts. This topic is a rapidly emerging field and it will be important to update the review regularly and to assess whether the categories of use cases identified here will be the same for new and emerging technologies.

## 5. Conclusion

NLP has made significant progress in information extraction and classification, which is conducive to automating and improving efficiency in clinical care processes.

Chatbots, sentiment analysis and decision support also have considerable potential benefits to improve patient-facing communication and support. However, there are currently limitations to implementing NLP in cancer care, including performance issues (accuracy) and a lack of evaluation in clinical practice. We recommend that researchers and practitioners work closely together to address existing issues and challenges.

## Supporting information

**S1 File. Keywords and search strategies.** This file records the keywords and search strategies for literature searches.

**S2 Table. Result extraction form.** This file records the detailed information that includes the title, first author, year of publication, scope of the paper and the problem addressed, NLP technology used, results, strengths and limitations.

**S3 File. Protocol for The Role of Natural Language Processing in Cancer Care-A systematic scoping review with narrative synthesis**.

**S4 Table. PRISMA checklist.**

## Supporting information

Supplemental File 1 Keywords and search strategies

Supplemental File 2 Result extraction Table

Supplemental File 3 Research Protocol

Supplemental File 4 PRISMA checklist

## Data Availability

All data produced are available online at the databases listed in the paper.

