## Supplemental File 1 Keywords and search strategies for "The role of natural language processing in cancer care: a systematic scoping review with narrative synthesis"

S1 Keywords and search update

The general syntax is to search single terms separately. Truncation and Wildcards will combine keywords as sub-searches and use Boolean operators to connect all results. Keywords and operators will be slightly modified for various databases or information sources. Use OR to connect the groups divided into two fields of computer and medicine and use AND to get the literature that includes terms from both fields.

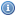
[<https://ovidsp.dc1.ovid.com/ovid-b/ovidweb.cgi?&S=JHBDFPODLIACNCGMKPMJLFEMMKCNAA00&Database+Field+Guide=26>](https://ovidsp.dc1.ovid.com/ovid-b/ovidweb.cgi?&S=JHBDFPODLIACNCGMKPMJLFEMMKCNAA00&Database+Field+Guide=26) **Embase Classic+Embase** 1947 to 2023 January 25**,** 
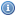
[<https://ovidsp.dc1.ovid.com/ovid-b/ovidweb.cgi?&S=JHBDFPODLIACNCGMKPMJLFEMMKCNAA00&Database+Field+Guide=29>](https://ovidsp.dc1.ovid.com/ovid-b/ovidweb.cgi?&S=JHBDFPODLIACNCGMKPMJLFEMMKCNAA00&Database+Field+Guide=29) **Ovid MEDLINE(R) and Epub Ahead of Print, In-Process, In-Data-Review & Other Non-Indexed Citations and Daily** 1946 to January 25, 2023

\# ▲	Searches	Results	Type

1	NLP {Including Related Terms}	8053	Basic

2	exp natural language processing/	15095	Advanced

3	NLG.mp. [mp=ti, ab, hw, tn, ot, dm, mf, dv, kf, fx, dq, bt, nm, ox, px, rx, ui, sy]	609 Advanced

4	natural language generat*.mp. [mp=ti, ab, hw, tn, ot, dm, mf, dv, kf, fx, dq, bt, nm, ox, px, rx, ui, sy]	156	Advanced

5	chatbot? {Including Related Terms}	1080	Basic

6	machine transla*.mp. [mp=ti, ab, hw, tn, ot, dm, mf, dv, kf, fx, dq, bt, nm, ox, px, rx, ui, sy]	559	Advanced

7	language model*.mp. [mp=ti, ab, hw, tn, ot, dm, mf, dv, kf, fx, dq, bt, nm, ox, px, rx, ui, sy]	2111	Advanced

8	exp information retrieval/	262402	Advanced

9	information extraction.mp. [mp=ti, ab, hw, tn, ot, dm, mf, dv, kf, fx, dq, bt, nm, ox, px, rx, ui, sy]	3590	Advanced

10	exp sentiment analysis/	469	Advanced

11	knowledge graph.mp. [mp=ti, ab, hw, tn, ot, dm, mf, dv, kf, fx, dq, bt, nm, ox, px, rx, ui, sy]	1240	Advanced

12	natural language understand*.mp. [mp=ti, ab, hw, tn, ot, dm, mf, dv, kf, fx, dq, bt, nm, ox, px, rx, ui, sy]	246	Advanced

13	dialogue manage*.mp. [mp=ti, ab, hw, tn, ot, dm, mf, dv, kf, fx, dq, bt, nm, ox, px, rx, ui, sy]	47	Advanced

14	automatic summar*.mp. [mp=ti, ab, hw, tn, ot, dm, mf, dv, kf, fx, dq, bt, nm, ox, px, rx, ui, sy]	98	Advanced

15	text classif*.mp. [mp=ti, ab, hw, tn, ot, dm, mf, dv, kf, fx, dq, bt, nm, ox, px, rx, ui, sy]	1316	Advanced

16	1 or 2 or 3 or 4 or 5 or 6 or 7 or 8 or 9 or 10 or 11 or 12 or 13 or 14 or 15	285035	Advanced

17	cancer care {Including Related Terms}	7349	Basic

18	aftercare {Including Related Terms}	13260	Basic

19	self management {Including Related Terms}	12065	Basic

20	Neoplasm {Including Related Terms}	110325	Basic

21	exp self care/	161143	Advanced

22	exp patient education/	211532	Advanced

23	cancer surviv* {Including Related Terms}	17441	Basic

24	cancer screen* {Including Related Terms}	18305	Basic

25	patient? monitor* {Including Related Terms}	16971	Basic

26	patient? assess* {Including Related Terms}	4916	Basic

27	clinic note?.mp. [mp=ti, ab, hw, tn, ot, dm, mf, dv, kf, fx, dq, bt, nm, ox, px, rx, ui, sy]	1351	Advanced

28 patient? feedback.mp. [mp=ti, ab, hw, tn, ot, dm, mf, dv, kf, fx, dq, bt, nm, ox, px, rx, ui, sy]	4343	Advanced

29	cancer treat*.mp. [mp=ti, ab, hw, tn, ot, dm, mf, dv, kf, fx, dq, bt, nm, ox, px, rx, ui, sy]	224579	Advanced

30 17 or 18 or 19 or 20 or 21 or 22 or 23 or 24 or 25 or 26 or 27 or 28 or 29	761740	Advanced

31	16 and 30	3051	Advanced

32	limit 31 to yr="2013 -Current"	1821	Advanced

33	remove duplicates from 32	1691	Advanced

**IEEE**

("Abstract":nlp OR "Abstract":"natural language processing" OR "Abstract":"natural language generation" OR "Abstract":"nlg" OR "Abstract":"nlu" OR "Abstract":"natural language understand*" OR "Abstract":"information retrieval" OR "Abstract":"sentiment analy*" OR "Abstract":"automatic summar*" OR "Abstract":"text classif*" OR "Abstract":chatbot OR "Abstract":"machine translat*" OR "Abstract":"machine reading comprehension" OR "Abstract":"dialogue manage*" OR "Abstract":"dialogue system" OR "Abstract":"knowledge graph" OR "Abstract":NER OR "Abstract":"information extra*" OR "Abstract":"text AI" OR "Abstract":"semantic analy*" OR "Abstract":"syntactic parsing" OR "Abstract":"word embedding") AND ("Full Text Only":"patient? feedback" OR "Full Text Only":"clinic note?" OR "Full Text Only":"patient? assess*" OR "Full Text Only":"patient? monitor*" OR "Full Text Only":"cancer screen*" OR "Full Text Only":"cancer surviv*" OR "Full Text Only":"patient? educa*" OR "Full Text Only":"cancer care" OR "Full Text Only":"self?management" OR "Full Text Only":aftercare OR "Full Text Only":neoplasm OR "Full Text Only":"self?care" OR "Full Text Only":"cancer diagnos*" OR "Full Text Only":"cancer treatment" OR "Full Text Only":tumor OR "Full Text Only":"cancer treat*") NOT ("Document Title":"image analy*" OR "Document Title":"image recogni*" OR "Document Title":"IoT")2013-2023

430

328 after duplicate

**Web of science**

(ALL=(nlp OR "natural language processing" OR "natural language generation" OR "nlg" OR "nlu" OR "natural language understand*" OR "information retrieval" OR "sentiment analy*" OR "automatic summar*" OR "text classif*" OR chatbot OR "machine translat*" OR "Machine Reading Comprehension" OR "dialogue manage*" OR "dialogue system" OR "knowledge graph" OR "NER" OR "information extra*" OR "text AI" OR "semantic analy*" OR "syntactic parsing" OR "word embedding")) AND ALL=("patient? feedback" OR "clinic note?" OR "patient? assess*" OR "patient? monitor*" OR "cancer screen*" OR "cancer surviv*" OR "patient? educa*" OR "cancer care" OR "self?management" OR aftercare OR neoplasm OR "self?care" OR "cancer diagnos*" OR "cancer treatment" OR tumor OR "cancer treat*")	2013-2023	1119

**ACM**

901 Results for: [[All: nlp] OR [All: "natural language processing"] OR [All: "natural language generation"] OR [All: "nlg"] OR [All: "nlu"] OR [All: "natural language understand*"] OR [All: "information retrieval"] OR [All: "sentiment analy*"] OR [All: "automatic summar*"] OR [All: "text classif*"] OR [All: chatbot] OR [All: "machine translat*"] OR [All: "machine reading comprehension"] OR [All: "dialogue manage*"] OR [All: "dialogue system"] OR [All: "knowledge graph"] OR [All: ner] OR [All: "information extra*"] OR [All: "text ai"] OR [All: "semantic analy*"] OR [All: "syntactic parsing"] OR [All: "word embedding"]] AND [[All: "patient? feedback"] OR [All: "clinic note?"] OR [All: "patient? assess*"] OR [All: "patient? monitor*"] OR [All: "cancer screen*"] OR [All: "cancer surviv*"] OR [All: "patient? educa*"] OR [All: "cancer care"] OR [All: "self?management"] OR [All: aftercare] OR [All: neoplasm] OR [All: "self?care"] OR [All: "cancer diagnos*"] OR [All: "cancer treatment"] OR [All: tumor] OR [All: "cancer treat*"]] AND [E-Publication Date: (01/01/2013 TO 12/31/2023)]

**ACL Anthology:**

"patient? feedback" OR "clinic note?" OR "patient? assess*" OR "patient? monitor*" OR "cancer screen*" OR "cancer surviv*" OR "patient? educa*" OR "cancer care" OR "self?management" OR aftercare OR neoplasm OR "self?care" OR "cancer diagnos*" OR "cancer treatment" OR tumor	**927**

262

Query:

nlp OR "natural language processing" OR "natural language generation" OR "nlg" OR "nlu" OR "natural language understand*" OR "information retrieval" OR "sentiment analy*" OR "automatic summar*" OR "text classif*" OR chatbot OR "machine translat*" OR "machine reading comprehension" OR "dialogue manage*" OR "dialogue system" OR "knowledge graph" OR NER OR "information extra*" OR "text AI" OR "semantic analy*" OR "syntactic parsing" OR "word embedding"

AND

"patient? feedback" OR "clinic note?" OR "patient? assess*" OR "patient? monitor*" OR "cancer screen*" OR "cancer surviv*" OR "patient? educa*" OR "cancer care" OR "self?management" OR aftercare OR neoplasm OR "self?care" OR "cancer diagnos*" OR "cancer treatment" OR tumor OR "cancer treat*"
