## Supplemental File 2 Result extraction Table for "The role of natural language processing in cancer care: a systematic scoping review with narrative synthesis"

S2 Table Result extraction form

| **Results extraction form** | | | | |
| --- | --- | --- | --- | --- |
| **Title, first author and year of publication** | **Scope of the paper and the problem addressed** | **NLP Technology Used** | **Results** | **Strength and limitations** |
| **Systems to support patient education and self-management (including emotional support, and self-management of distress)** | | | | |
| **Educating patients** |  |  |  |  |
| Development and Early Feasibility of Chatbots for Educating Patients with Lung Cancer and Their Caregivers in Japan: Mixed Methods Study; Yuki Kataoka; JMIR Cancer; 2021 | Developed and evaluated the early feasibility of a chatbot designed to improve the knowledge of symptom management among patients with lung cancer in Japan and their caregivers.By classified user questions and offering specific response. | Bot Designer (LINE Corp) for design and bot+Google Cloud's Dialogflow for generating response | 1)14 medical staff participated in the α test: 71 questions to the chatbot during this period. 11 (15%) questions did not match the author's question categories.  2)11 patients and 1 caregiver participated in the β test: The participants were asked 60 questions, of which 8 (13%) did not match with the appropriate question categories. 3 (5%) questions based on daily conversation did not have corresponding responses.  3)Satisfaction score was 2.7 (SD 0.5) points out of 5 | Limitations: 1)8 (13%) questions did not match suitably with responses in phase 5, and a patient complained of not receiving appropriate responses. It is necessary to add educational categories and responses through further discussion.  2)This was a single-centre study, and further studies are needed to evaluate the applicability of the findings in other hospitals. |
| A Personalized Data-to-Text Support Tool for Cancer Patients; Saar Hommes; INLG; 2019 | Developed a data-to-text system for cancer patients, providing information on quality of life implications after treatment | Rule based PASS data-to-text system | Both clinical physicians agreed that examples illustrating how quality of life impacts patients' lives were very useful. However, one physician noted that not all quality of life indicators like percentage of problems might happen should be conveyed in the same manner. Additionally, one physician pointed out that predicting how treatment will affect a patient's financial situation depends on many external factors, should consider more complex methods. | 1.Lack of evaluation of a batch set of data,to check accuracy or user satisfactory |
| Creating and Evaluating Chatbots as Eligibility Assistants for Clinical Trials: An Active Deep Learning Approach towards User-centered Classification;CHING-HUA CHUAN; ACM Transactions on Computing for Healthcare; 2020 | Developed a chatbot with a backend machine learning module to help users understand eligibility criteria for clinical trials. | Chatbot(conversation manager)+criteria classifier+active learning(uncertainty cluster sampling and label propagation with simulated annealing) 1. Identifying user intent, 2. Classifying clinical trial criteria, and 3. Generating conversations using NLP methods and a chatbot (conversation manager) with a criteria classifier and active learning support | Accuracy of classifying clinical trial was higher than 90%. When comparing web-bot and chatbot, the ratings on perceived usability and dialogue were significantly higher in the chatbot condition than the web-bot condition. In the chatbot-only condition, 97% asked the chatbot questions about medical terms, and only one participant used the search engine instead of the chatbot. |  |
| HELPeR: An Interactive Recommender System for Ovarian Cancer Patients and Caregivers; Behnam Rahdari; ACM RecSys Conference on Recommender Systems; 2022 | Built a knowledge-adaptive interactive recommender system for ovarian cancer patients and their caregivers to recommend relevant articles and topics. | 1.HELPeR Library Crawler 2.HELPeR Library Organizer:sectioning,topic modeling, key-phrase extraction, identifying detailed information such as required knowledge level  3.HRLPeR Patient information Processor  4.HRLPeR Recommender Engine HELPeR Patient Information Processor moudules, use Cypher query lanauage for Neo4j database to generate the search and recommendation results. | No evaluation | The paper was exploratory and investigated a potential use case for the technology. The technology was not evaluated with target users. |
| **Analysing patients’ emotions** |  |  |  |  |
| Empathic conversational agents for real-time monitoring and co-facilitation of patient-centered healthcare;Achini Adikari;Future Generation Computer Systems;2021 | Monitored (what? Online forum conversations?) in real-time to predict patients’ subsequent emotions (positive or negative) in their next online post. to ……. | 1.Word2vec for emotion sequence extraction  2. Markov chains for emotion state transitions  3. Second-order Markov model for emotion prediction  4.Feature engineering+ TFIDF+ensemble machine learning models presictions for group emotion prediction  5. Rule-based emotional message generation  6. All in a chatbot framework | It was observed that in 79% instances, the model was able to correctly predict the next emotion as positive or negative, and in 63% instances, the correct emotion out of all emotions was predicted. | Emotion extraction can be improved by addressing the current limitations related to detecting ambiguous expressions, idioms and indirect emotion statements. The author mentioned advanced NLP such as transformer-based language models, will be used to improve the contextual understanding of the patient conversations. |
| Natural Language Processing–Based Virtual Cofacilitator for Online Cancer Support Groups: Protocol for an Algorithm Development and Validation Study; Yvonne W Leung; JMIR RESEARCH PROTOCOLS; 2021 | Developed and evaluated an artificial intelligence–based cofacilitator (AICF) prototype to track and monitor online support group participants’ distress through real-time analysis of text-based messages posted during synchronous sessions. | Applied the framework called PRIME, including emotion extraction, relevant terms dictionary, information mapping and validation by human experts. | When compared with the human annotation of distress levels, the AICF was able to correctly identify specific types and intensity of emotion from the 8 participants in 10 sessions for 78% of instances for low distress, 63% for moderate distress, and 85% for high distress in tech. The baseline performance for distress classification was 72% agreement with the annotators with advanced training in clinical psychology. | One limitation is that the AICF relies on human annotated training data and human scoring for performance evaluation. |
| Providing Care Beyond Therapy Sessions With a Natural Language Processing–Based Recommender System That Identifies Cancer Patients Who Experience Psychosocial Challenges and Provides Self-care Support: Pilot Study; Yvonne W Leung; JMIR CANCER; 2022 | Evaluate the AICF prototype above on its precision and recall in recommending resources to cancer Online Support Group (OSG) members | Took the PRIME framework and extended the functionality to generate notifications to patients | F1 score reach 0.88,19 (76%) participants rated the stress recommendation as "useful" | These resources were selected for their relevance to assist OSG participants in dealing with the psychosocial challenges of living with different cancers. However, such resources could also be seen as too generic by participants and insufficient to meet their needs for a specific cancer. This may partially explain the fact that only 52% of the participants accessed a recommended resource. |
| **Systems to improve efficiency in clinical care by summarizing data, extracting data, categorizing data, and supporting record keeping** | | | | |
| **Extract information from medical report** |  |  |  |  |
| A Question-and-Answer System to Extract Data From Free-Text Oncological Pathology Reports (CancerBERT Network): Development Study; Joseph Ross Mitchell;JMIR; 2022 | Developed a BERT-based system to automatically extract detailed tumor site and histology information from free-text oncological pathology reports. | Initialized weighted from BERT based model and finetuned with electronic pathology reports. Connects caBERT A,B,C,use for site and histology question and answering, ICD-O-3 histology code classification. | Accuracy:  caBERT A Trained site&histology Q&A:79.99% caBERT B Trained site classification: 73.84%, (93.28% top5) caBERT C Trained histology classification: 85.29%,(96.27% top5) | Strength: This study included more cancer categories and obtained higher accuracy scores. Limitations: 1. Top 1 accuracy is not good enough to use. 2. The Q&A task expect a single answer to each question asked of each input may different from the real use case. |
| Mining Electronic Health Records to Extract Patient-Centered Outcomes Following Prostate Cancer Treatment;Tina Hernandez-Boussard;AMIA Annu Symp Proc;2017 | Developed and validated an NLP pipeline system to detect important patient-centered outcomes (PCOs) as interpreted and documented by clinicians in dictated notes for male patients undergoing treatment for localized prostate cancer at an academic medical center | Using GATE software for data processing (1) an ANNIE module to extract PCO mentions from narrative texts, (2) a ConText module to determine the semantic context of the PCO mentions, and (3) a JAPE module to annotate the PCO mentions in the text. | The system achieves average F1 scores of 90% | First, the algorithms have been developed and tested in a single academic center. A second limitation is that the system only reports what the clinician documents and does not capture patient-reported outcomes. |
| NLP Automation to Read Radiological Reports to Detect the Stage of Cancer Among Lung Cancer Patients;Er. Khushbu Gupta; Workshop on Widening NLP; 2019 | Create medical reports annotations in clinical practice system automatically where the computer can read radiological reports for oncologists and mark the staging of lung cancer. | 1.Rule-based Clinical Information Extraction, Structured Data, Verification with manual rules of TNM staging, Get Staging Labels. 2. LSTM based extraction method | 1.heuristic approach, accuracy of the system was achieved up to 59% for T-staging, 36% for N-staging and 41% for M-staging 2.implementing LSTM, the system could bring up to 85% of accuracy for the collective TNM staging. | Limitations:Only 23 reports were annotated for evaluation. May not be reliable |
| A Natural Language Processing Program Effectively Extracts Key Pathologic Findings from Radical Prostatectomy Reports; Brian J. Kim; JOURNAL OF ENDOUROLOGY; 2014 | The objective of this study was to evaluate the performance of an internally developed NLP program in extracting select pathologic ﬁndings from radical prostatectomy specimen reports in the EMR. | Applied a system called KPSC Clinical Information Extraction System to extract key variables from prostatectomy reports in the EMR within the healthcare system | NLP demonstrated 100% accuracy for identifying the Gleason grade, presence of a tertiary Gleason pattern, SVI, ALI, and ECE. It also demonstrated near-perfect accuracy for extracting histologic subtype (99.0%), PNI (98.9%), TNM stage (98.0%), SMS (97.0%), and dominant tumor size (95.7%). The overall accuracy of NLP was 98.7%. | The author noted it is challenging to determining the size of primary tumor. This due to differences in the language used to describe tumors, such as "mass," "lesion," and "nodule." Furthermore, many structures beyond the tumor itself are often measured and described in a given report, leading to possible confusion in the software. NLP may succeed when extracting more explicit terms such as Gleason sum, tertiary pattern, and T-phase. |
| **Restructuring medical reports** |  |  |  |  |
| Automatic Population of Structured Reports from Narrative Pathology Reports; YING OU; HIKM 2014 Seventh Australasian Workshop on Health Informatics and Knowledge Management | Used natural language processing to extract pertinent information from free-text pathology reports to automatically populate structured reports. | Identified medical entities with supervised machine learning approaches CRF, and then applied rule-based methods to construct the output for the report population with the correct entities. | The evaluation on the training set shows that the system performance can be improved by about 8.7% through rule refinement. The overall micro-average precision, recall, and F-score for the end-to-end evaluation on the test set are 89.44%, 80.60%, and 84.79%, respectively. No structured reports evaluation. | Limitation: The study was limited to a specific clinical domain and genre, so it might have a lack of generalizability or portability. Not evaluated beyond a training dataset |
| DeepPhe: A Natural Language Processing System for Extracting Cancer Phenotypes from Clinical Records; Guergana K. Savova; AACR American Association for Cancer Research; 2017 | Automated extraction of detailed phenotype information (tumor morphology, laboratory results etc.,) from electronic medical records of cancer patients | Level 1: a cTAKES system extract mentions of system basic concepts  Level 2: Fast Health Interoperability model summarizes mentions from a single doucument into a template Level 3&4: classify a given document into episode categories and make summarization based on template | The DeepPhe system's performance was compared to human expert abstracted information. Human expert agreement ranged from 0.46 to 1.00, while system agreement ranged from 0.20 to 0.96. The study used a metric based on sensitivity and positive predictive value to measure agreement, showing that DeepPhe performed similarly to human experts. | Strength:  The paper showed the potential to save time and costs associated with manual cancer phenotype abstraction  Limitation: Evaluation not follow specific technology or precise medical methods. |
| Effective Natural Language Processing and Interpretable Machine Learning for Structuring CT Liver-Tumor Reports; YI-HSUAN CHUANG; IEEE Access; 2022 | Developed a report structuring method by integrating NLP and interpretable machine learning for further disease risk assessment, disease recognition and treatment recommendation | Off-line training: including data processing, N-gram keywords extraction, determinations of feature keywords, train multi-classifier (RF, SVM,LDA,NN) by keywords Online structuring: n-gram keywords extraction, generation of feature vectors and multi-labels classification. Predicted labels return to the form. | The optimal result used NN with one+bi+tri gram models and classifier accuracy achieved 0.90519 | Strengths: Performed better than bert based model Tbert,Sbert,Bbert,Cbert, and training time only needs 2.66 seconds.  Limitations:1. The parameters in the proposed method were approximated for the experimental data. 2. The ﬁnal structure was deﬁned by a few radiologists in Taiwan. |
| **Classifying data from diagnostic and clinical reports** |  |  |  |  |
| Improving natural language information extraction from cancer pathology reports using transfer learning and zero shot string similarity; Briton Park; JAMIA Open; 2021 | Classified tumor attributes from pathology reports given minimal labeled examples | Hierarchical cancer to cancer transfer (HCTC) and zero-shot string similarity (ZSS) methods were designed to exploit shared information between cancers and auxiliary class features | The model maximum F1 macro score achieved was 0.5, and the micro score was 0.86. | The author found that using pre-trained word embeddings performed worse in measuring similarity compared to string-based methods but still believe that leverage models pretrained on large corpora for language modeling task is promising. |
| Knowledge-Powered Deep Breast Tumor Classiﬁcation With Multiple Medical Reports; Dehua Chen; IEEE/ACM TRANSACTIONS ON COMPUTATIONAL BIOLOGY AND BIOINFORMATICS;2021 | Classified breast tumors using multiple medical reports such as B-ultrasound, Mammography (X-ray) and Nuclear Magnetic Resonance Imaging (MRI). | Model ﬁrst uses Hierarchical Attention Bidirectional Recurrent Neural Networks (HA-BiRNNs) to encode the syntax-aware representation of medical reports in a hierarchical way. Secondly, the model obtains the semantic information relevant to the medical reports from the clinical domain semantic tree and encodes the semantic representation of medical reports by using Tree Structured Recurrent Neural Network with gated recursive units (Tree-GRUs). Finally, the author classify breast tumors by combining both the syntax and semantic representations of medical reports. | The KDBTC method of applying syntax-aware medical report encoding module uses HABRNN and the semantic encoding module uses Tree-GRUs achieve the best performance of 0.8854 accuracy and 0.9070 F1 score | Considering the syntax information, the author mentioned using attention mechanisms is better than without using attention mechanism, and the performance of using bidirectional recurrent neural networks is better than using recurrent neural networks. And the result of classiﬁcation using both the syntax and semantic information is better than only syntax information |
| Natural Language Processing to Ascertain Cancer Outcomes From Medical Oncologist Notes; Kenneth L.; JCO Clinical Cancer Informatics; 2020 | Use neural network-based NLP to extract structured clinical outcomes from unstructured medical oncologist notes. | 1. Rule based classifier to recognize the assessment/plan for a given note, key phrase was then removed from each note, a RNN was trained to predict whether each remaining word in each note was part of the plan  2. CNN trained to predict the labels applied by a human curator to assessment/plan text for the following 3 binary outcomes: whether cancer was present; whether cancer was progressing or worsening; and whether cancer was responding or improving | AUROCs of 0.94 for the any-cancer classifier outcome, 0.86 for the progression classifier outcome, and 0.90 for the response outcome. | Limitations: 1.Studies only focus on lung cancer patients 2.oncologists sometimes uncertain about whether a patient’s cancer is responding or progressing at the time that a clinical note is ﬁled. 3.cases in which human curators were unsure whether to indicate an outcome as present or indeterminate might have yielded noisy target labels(wrong samples). |
| Phenotyping severity of patient-centered outcomes using clinical notes: A prostate cancer use case; Selen Bozkurt; Learn Health Syst; 2020 | Used natural language processing (NLP) to phenotyping patients with Urinary incontinence (UI) to classify their incontinence symptom into severity subtypes, so that these patients could be offered more tailored advice/treatment. | Using a hybrid NLP pipeline that combines rule-based and deep learning methodologies, classified positive UI cases as mild, moderate, and severe by mining clinical notes. | The system classified positive urinary incontinence I case as mild, moderate, and severe, with F1 scores of 0.76, 0.81, and 0.71 respectively. | Limitations:  Not considering do clinicians used UI rating to improve treatment |
| **Support record keeping** |  |  |  |  |
| Weakly supervised natural language processing for assessing patient-centered outcome following prostate cancer treatment; Imon Banerjee; Scientific Reports; 2021 | Used an NLP pipeline to assess the presence, absence, or risk discussion urinary incontinence (UI) and bowel dysfunction (BD) following prostate cancer treatment. | A weighted function of neural word embedding was used to create a sentence-level vector representation of relevant expressions extracted from the clinical notes. Sentence vectors were used as input for a multinomial logistic model, with output being either presence, absence or risk discussion of UI/BD. The classiﬁer was trained based on automated sentence annotation depending only on domain-speciﬁc dictionaries (weak supervision). | Average classifier F1 score of 0.86 for the sentence-level, three-tier classiﬁcation task (presence/absence/risk) in both UI and BD | Limitations: 1. Assessing outcomes need bigger dataset 2.Domain-specific dictionary is limited 3.Lacks sensitivity for word order which limits the ability of learning long-term and rotated scope of negex terms. |
| **Systems to support prevention and early detection of patient problems or cancer recurrence through risk stratification** | | | | |
| **Risk assessment and follow-up care monitoring** |  |  |  |  |
| A Smartphone App Framework for Segmented Cancer Care Coordination;Tiancheng He;(BHI) IEEE-EMBS International Conference on Biomedical and Health Informatics ;2016 | Developed a smartphone app that provides cancer risk assessment and follow-up care monitoring services | The mHealth application framework includes three functional modules: a Bayesian model-based natural language processing module for extracting relevant information from free text or medical reports; a cancer risk calculator, using support vector machine classification, based on the extracted information Assessing the medical risk of cancer patients; and a healthcare monitor that provides timely care coordination for high-risk cancer patients. | (1) Medical Information Retrieval:The average accuracy of all parameters was 97%. (2) Medical risk assessment: To cover all high-risk patients, the sensitivity of the test results was close to 100%, but the specificity of the test results was only 70%, which means that a small number of low-risk patients are included in daily health care monitoring.  (3) Health monitoring: During the evaluation period, the doctor communicated with each patient at least once a day. No result. | Limitation: The health monitor was not developed. |
| **Cancer recurrence prediction** |  |  |  |  |
| Predictive models for colorectal cancer recurrence using multi-modal healthcare data; Danliang Ho; ACM CHIL; 2021 | Used heterogeneous healthcare data to predict colorectal cancer recurrence | 1) a Transformer model carefully modified to extract high-quality features from time-series data, and 2) a Multi-Layered Perceptron (MLP) that learns tabular data features, followed by feature integration and classification for prediction of recurrence. | AUROC score of the model was 0.95, as well as precision, sensitivity and specificity scores of 0.83, 0.80 and 0.96 respectively, surpassing the performance of all-known published results based on CEA, as well as most commercially available diagnostic assays. | Limitation:1.Limited in the specific areas. 2.It will also increase clinical relevance if the model also possesses the ability to proactively forecast recurrence months before onset. The author note that similar to existing multi-modal architectures, the model lacks interpretability; quantifying feature contributions at the individual and modality level may improve the trustworthiness of the model by shedding light on whether the model is outputting predictions based on reasonable explanations. Lastly, regarding measure of generalisation performance, due to time constraints the authors could only perform 5 bootstrap evaluations; note that 50-200 bootstrap samples were recommended for obtaining reliable estimates of uncertainty and hope to do so given more time. |
| Weakly supervised temporal model for prediction of breast cancer distant recurrence; Josh Sanyal; Scientific Reports;2021 | Predicted breast cancer distant recurrence using free-text clinic notes by a weak-supervision framework | 1)t-SNE for word embedding 2)2 wea-supervision approach, one train model with manually-curated data and NLP-generated labels with optimal sensitivity and specificity, one —trained with manually-curated data and the NLP-generated labels with high sensitivity,against a traditional prediction model 3)temporal LSTM model to predict recurrence | achieved 0.94 AUROC of the framework |  |
| **Short-term life expectancy prediction** |  |  |  |  |
| Probabilistic Prognostic Estimates of Survival in Metastatic Cancer Patients (PPES-Met) Utilizing Free-Text Clinical Narratives; Imon Banerjee; Scientific Reports; 2018 | Estimated short-term life expectancy (>3 months) of the patients by analyzing free-text clinical notes in the electronic medical record, while maintaining the temporal visit sequence. | 1. Develop a hybrid pipeline that combines semantic data mining with neural embedding for creating context-aware dense vector representation of the multiple types of free-text clinical notes.  2. Proposed an efficient deep prognosis model that takes as input the context-aware vectorized representation of sequential clinic notes and outputs a probability of short-term life expectancy estimate (>3 months).  3. Incorporated an interactive visualization method for improving physician understanding of the basis for the model’s predictions. | ROC 0.89 of prognosis model | 1.First, the author reported many patients in the current training dataset lost follow-up  2.Second, the time points were not equally spaced, and a single day may contribute multiple data points.  3.the model was trained using single institutional data that contains biases regrading syntactic style of clinical narratives, patient populations, treatment planing.  4.the longitudinal survival cureve for identifying the core finding of the visit notes via semantic data mapping and context analysis is not statistically evaluated in this study, but only presented as a tool for understanding basis of model prediction. |
| **Symptoms to improve cancer treatment by supporting clinicians to make evidence-based treatment decisions** | | | | |
| Comparison of an oncology clinical decision-support system’s recommendations with actual treatment decisions;Suthida Suwanvecho;Journal of the American Medical Informatics Association;2021 | Treatments selected by clinicians in BIH (Bumrungrad International Hospital) were paired with therapeutic options presented by the CDSS and coded to mask the origin of options presented. | WfO(Watson for Oncology) system that provides treatment recommendations for a given case using relevent attributes | Nearly 60% (187) of 313 treatment pairs for breast, lung, colon, and rectal cancers were identical or equally acceptable, with 70% (219) of WfO therapeutic options identical to, or acceptable alternatives to, BIH therapy. In 30% of cases (94), 1 or both treatment options were rated as unacceptable. Of 32 cases where both WfO and BIH options were acceptable, WfO was preferred in 18 cases and BIH in 14 cases. Colorectal cancers exhibited the highest proportion of identical or equally acceptable treatments; stage IV cancers demonstrated the lowest. This study demonstrates that a system designed in the US to support, rather than replace, cancertreating clinicians provides therapeutic options which are generally consistent with recommendations from oncologists outside the US. | These findings illustrate the fact that humans in practice do not always choose the best course of treatment, identifying a gap where CDSS could improve performance. |
| Extracting Features from Online Forums to Meet Social Needs of Breast Cancer Patients; Maitreyi Mokashi; ACM SIGCAS Conference on Computing and Sustainable Societies;2020 | Provide a platform for health care providers to understand and acknowledge the “personal and social issues” a patient might encounter at the current or future state of the disease. Additionally, the physician may get an estimate of what issues the patient might face due to the new treatment plan. | 1.Bipartite network – BiNe +random walk method in Node2Vec for embedding 2.For detecting the communities the authors use k-means clustering and learn the clusters using Expectation Maximization (EM) algorithm. | The Patient-Topic Network coherence, measured by Normalized Pointwise Mutual Information (NPMI) of the clusters, was 0.481, and the Precision and Recall were 0.643 and 0.588, |  |
| Machine learning to support social media empowered patients in cancer care and cancer treatment decisions; Daswin De Silva; PLoS ONE; 2018 | Investigate the impact of online social influences on the intimate decision scenario of selecting a treatment type, recovery after treatment, side effects and emotions expressed over time, using prostate cancer as a model. | Patient Reported Information Multidimensional Exploration (PRIME) framework for automated investigation of patient behaviours, clinical factors and patient emotions, across the temporalities of diagnosis, treatment and recovery  Stage1: collocate OSG conversations by patients  S2: External NLP technology for information extraction  S3: Rules and clinical ontologies for information enrichment and regulat expression classifier for composition  S4: Rule mining to infer decision-making behaviour and decision factors  S5: Vocabulary based extraction of patient reported side effects  S6-7: Emotion extraction  (1)Word2vec and seed thesaurus for extract semantically similar terms  (2)Domian specific emotion terms curated by experts and Intensity modifier dictionary and pre-processed OSG posts are used for emotion analysis to generate emotion intensity vectors  Output: Decision factors at t0, patient reported side effect t0 to t12, emotion expressions from t-3 to t12(tn) | Analysis of 1.the composition of each group in terms of volume, age, grading of cancer (using Gleason score) and modality of treatment.  2.the diversity of decision factors, ranging from clinical skills to financial concerns  3.comprehensive analysis of distinct patient emotions, expressed over timefrom pre-decision to recovery  4.side effects, with a higher percentage of Shared group reporting all side effects than the other two groups. |  |
| Natural language processing of German clinical colorectal cancer notes for guideline-based treatment evaluation; Matthias Becker; International Journal of Medical Informatics; 2019 | Identify speciﬁc guideline-based patient information and to annotate it with Uniﬁed Medical Language System concepts for manual evaluation by a physician. | The system was mainly based on string manipulation and the Uniﬁed Medical Language System (UMLS) dictionary and was evaluated using data obtained from German EHR clinical notes annotated by a physician. | The average system precision value across all concepts relevant to treatment decisions for patients with known cancer diagnoses achieved an F1 score of 81.81%. |  |
| Constructing Ontology-based Cancer Treatment Decision Support System with Case-Based Reasoning; Ying Shen; SmartCom 2017 Smart Computing and Communication | Disease Ontology (DO) that pertains to cancer’s clinical stages and their corresponding information components was utilized to improve the reasoning ability of a decision support system (DSS). The proposed DSS uses Case-Based Reasoning (CBR) to consider disease manifestations and provides physicians with treatment solutions from similar previous cases for reference. | Rule Based medical reasoning framework combined operation of DSS, CBR and ontology(method, cure, cancer) triplet information) can aide clinical decision-making.  To simplify the semantic representation of medicines, existing biomedical ontology was reused in DSS.D9 | the AUC of the DSS with ontology was 0.846, while without ontology was 0.73 |  |
| Precision Medicine Search for Paediatric Oncology;Bevan Koopman; ACM SIGIR 21: Proceedings of the 44th International ACM SIGIR Conference on Research and Development in Information Retrieval; 2021 | A search engine aimed to help clinicians find targeted treatments for children with cancer. | Entity extraction was done to capture three key types of information: (drugs, cancers, genes). Using BERN: a neural medical, named entity recognition tool. Result display was also based around these entity types, with an interactive, entity-based knowledge graph constructed, providing clinicians with a more interpretable alternative to a traditional SERP. | No evaluation | Strength: Provided a very targeted scope.  Limitations: Not verify the actual use effect |
