## Supplemental File 3 Research Protocol for "The role of natural language processing in cancer care: a systematic scoping review with narrative synthesis"

### Background

Cancer is the leading cause of premature death worldwide. Global Cancer Statistics 2020 [1] shows an estimated 19.3 million new cancer cases and nearly 10 million cancer deaths worldwide. Cancer care is a rapidly developing field that encompasses various processes, from screening and early detection, through treatment, [2] survivorship care, and end of life care. Natural language processing (NLP) is a promising tool that could be used to enhance the efficiency, treatment quality, and patient experience of cancer care.

NLP is a study that merges the fields of artificial intelligence and linguistics, primarily focusing on text recognition, understanding, and generation, with machine learning, deep learning, and linguistics as fundamental research [3]. NLP technologies have been increasingly used in industry to build chatbot assistants [4], extract meaningful information from big data [5], recommend articles or treatment suggestions based on user behavior and preference [6], and generate structured or simplified reports from long and complex documents [7].

In cancer care, NLP can be used to extract and analyze information from various sources, including electronic health records (EHRs), clinical notes, pathology reports, radiology reports, and social media data [8]. This data could aid cancer treatment and patient self-management and could help healthcare providers make treatment decisions. For example, chatbots and virtual assistants can inform patients about their condition, treatment options, and potential side effects. NLP has also been used to extract relevant information from pathology reports, such as tumor type, stage, and grade, which can inform treatment decisions [9].

This scoping review aims to explore the research and application of NLP in cancer treatment and patient self-management. Overall, the role of NLP in cancer care is rapidly evolving and holds great promise for improving the quality and efficiency of patient care. As more data becomes available and NLP technologies continue to advance, it is expected that NLP will become an increasingly valuable tool for cancer care providers. This review will summarise the ways in which NLP has been used to improve the management of cancer by individuals with cancer and/or their clinicians.

### Review Questions

The following are specific research questions.

1. What roles can text data provide in cancer patients' self-management?
2. What type of support can or has NLP technology provided to cancer patients and their clinicians?

3. What are the challenges or barriers to using NLP technology to improve patient outcomes in oncology (for example, technical, privacy, ethical, and implementation)?

**Main aims of the review**

Our aims for doing this scoping review are as follows:

1. Identify the current state of research on using NLP in cancer treatment, including the types of cancer studied, the NLP methods used, and the outcomes measured.
2. Assess the quality and limitations of the existing literature on NLP in cancer care, including gaps and areas for further research.
3. Identify potential benefits of NLP in cancer care, such as improved patient outcomes, better communication between healthcare providers and patients, and more efficient use of healthcare resources.
4. Assess the challenges and barriers to implementing NLP in cancer care, such as data privacy and security concerns, variability in clinical language, and limitations in the accuracy and reliability of NLP algorithms.
5. Recommendations for future research on NLP in cancer treatment include developing more advanced NLP methods, evaluating the effectiveness and safety of NLP-based interventions, and exploring NLP's ethical and legal implications in healthcare.

This scoping review will guide the effectiveness of future systematic reviews or functional developments aimed at helping cancer patients transition to more efficient and high-quality cancer care.

**Searches**

The literature will be identified by searching medical databases: MEDLINE and Embase, and the scientific and technical databases: Web of Science, IEEE, ACM, and NLP database ACL Anthology. This search strategy will ensure that the scope of information obtained is comprehensive.

Search terms are based on the topic and relative sub-tasks, and the Medical Subject Headings (MeSH) are used to get synonyms to broaden the scope. A medical research librarian was consulted for guidance on creating a search strategy. Truncation and Wildcards will combine keywords as sub-searches and use Boolean operators to connect all results. Keywords and operators will be slightly modified for various databases or information sources.

The general syntax is to search single terms separately. Use OR to connect the groups divided into two fields of computer and medicine and use AND to get the literature that includes terms from both filed. The publication time is from 2013 to the present (2023). Examples are as follows:

(NLP OR “automatic summar*” OR “sentiment analy*” OR “dialogue system*”) AND ("cancer care" OR "cancer treatment" OR "patient? educa*")

**Identifying relevant studies**

Article screening will be conducted in three stages using RefWorks, following the Fig 1 PRISMA flowchart[10], including 1) title screening, 2) abstract screening, and 3) full-text screening, with the selection process and filtering results recorded. After de-duplication, all the references will be divided equally among three reviewers with diverse backgrounds to screen the titles. The fourth reviewer will screen all the titles one by one. The four reviewers will meet after the title and abstract screening to discuss and resolve any conflicts. Full-text results will be obtained for all results that are judged to be potentially relevant and will also be dual-screened. The authors will meet for a second time to resolve any conflicts.

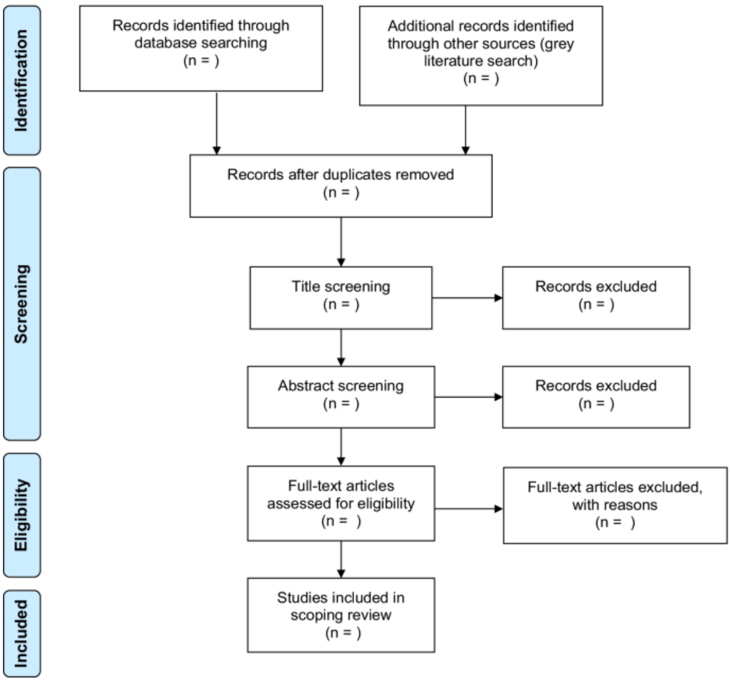


Figure 1 PRISMA flowchart for study selection[10]

**Inclusion Criteria**

In this review, we focus on the scope and potential benefits of NLP for cancer management. Studies will meet the following criteria.

1. Studies focus on applying NLP technologies to improve cancer management by patients, caregivers, or clinicians.
2. Studies focus on any cancer type and at any stage in the disease trajectory after diagnosis to survivorship or end-of-life care.

3. Studies include individuals of all ages – children and adult patients or that describe NLP systems designed to improve the clinical care of these patients.
4. Technologies that use patient self-reports to signpost to relevant educational resources or to directly deliver therapies or support, e.g., to help manage stress, mental health, or emotions.
5. Research articles in journals or conferences (including conference abstracts where full text/detailed descriptions are also available).
6. Studies that include original research including the following study types: clinical trials, development papers, and feasibility studies.
7. No article language or programming language restrictions will apply to the research.

**Exclusion Criteria**

1. Studies that do not focus on individuals with a confirmed cancer diagnosis, e.g., technologies to predict cancer risk, cancer prevalence, or to assess for potential symptoms of cancer. Death-related applications, such as mortality rate prediction, are not considered.
2. Papers that use cancer datasets for NLP research without focusing on improving cancer management by patients, caregivers, or clinicians.
3. Papers that use NLP to search research or medical databases such as PubMed.
4. Studies that do not involve both cancer care and NLP.
5. Studies for which full-text publications are not accessible or conference abstracts without associated full-text descriptions of the research.
6. Studies that do not include a medical or NLP evaluation of the technology/intervention.

**Data Extraction**

Once the studies have been selected, the next step is to extract the data from each study. The data extraction form is as follows

| # | **ITEMS** |
|---|---|
| 1 | Title |
| 2 | Name of the first author |
| 3 | Year of publication |
| 4 | Journal/Conference |
| 5 | System Objectives |
| 6 | Language of applications |
| 7 | Country of origin |
| 8 | Cancer type(s) included |
| 9 | Distribution of the target audiences |
| 10 | Ways in which NLP has been used to affect cancer management |
| 11 | NLP tasks or typical models |
| 12 | Dataset |

| | |
|---|---|
| 13 | Evaluation Methods |
| 14 | Key findings |

**Method of synthesis**

A scoping review approach was chosen because we wish to determine the breadth of existing research and our research questions cut across different disciplinary domains (including computing science and medicine).

Data will be analyzed descriptively, both quantitively and qualitatively. Data will be summarized, including the number of papers, the language of applications, country of origin, cancer types, system objectives, journal/ conference, distribution of the target audiences, NLP tasks and system architecture.

The studies will be categorized and summarized. For example, research will be categorized as being conducted primarily as a theoretical NLP/computing science research, or primarily as a practical health care application. The key tasks that the NLP system is performing, the aims and health domains covered by the system, results of any evaluation (e.g., usability, acceptability, ability to alter patient outcomes) evaluation methods will be described.

The NLP part will be analyzed from NLP tasks and methods, innovation elements, problem-solving points, and evaluation methods. First, data will be summarized on the ways in which NLP has assisted with cancer care, the nature of the NLP task and in which way NLP help to improve cancer. Second, whether the dataset is reliable and representative. Third, what innovations have been made in the algorithm and structure, and last whether the evaluation method is comprehensive or innovative.

From the perspective of cancer care, we will analyze 1) How many types of cancers are covered, how many countries, patient demographics/intended recipients of the technology 3) evaluation methods 4) findings of any evaluation including strengths and limitations of the technology, feasibility, acceptability, and any ethical or data security issues or concerns.

**Discussion**

This review will use transparent and replicable procedures to describe the scope of existing literature . This protocol outlines the data sources, search strategies, and data extraction methods. The scoping literature review aims to identify the application scenarios of NLP in the field of cancer treatment, analyze usability and limitations, and discuss how to improve patient care, increase efficiency, and reduce costs.

By conducting a thorough scoping literature review, we can better understand the opportunities and challenges associated with NLP in cancer treatment and provide insights that can inform the development of more effective and efficient healthcare NLP systems.
